# AEROSOL GENERATION DURING COUGHING - QUANTITATIVE DEFINITION FOR AEROSOL GENERATING PROCEDURES: OBSERVATIONAL STUDY

**DOI:** 10.1101/2021.08.24.21262520

**Authors:** Enni Sanmark, Lotta-Maria A.H. Oksanen, Noora Rantanen, Mari Lahelma, Veli-Jukka Anttila, Lasse Lehtonen, Antti-Pekka Hyvärinen, Ahmed Geneid

## Abstract

**Aim:** The purpose of the study was to determine aerosol exposure generated by coughing in operation room environments to create a quantitative limit value for high-risk aerosol-generating medical procedures.

**Background:** Coughing is known to produce a significant amount of aerosols and is thus commonly used as a best reference for high-risk aerosol-generation. Accordingly, procedures during which aerosol generation exceeds the amount of aerosol generated in instances of coughing are seen as high-risk aerosol generating procedures. However, no reliable quantitative values are available for high-risk aerosol-generation.

**Methods:** Coughing was measured from 37 healthy volunteers in the operating room environment. Aerosol particles generated during coughing within the size range of 0.3–10 µm were measured with Optical Particle Sizer from 40cm, 70cm, and 100cm distances. The distances reflected potential exposure distances where personnel are during surgeries.

**Results:** A total of 306 coughs were measured. Average aerosol concentration during coughing was 1.580 ± 13.774 particles/cm^3^ (range 0.000 – 195.528).

**Discussion:** The aerosol concentration measured in this study can be used as a limit for high-risk aerosol generation in the operating room environment when assessing the aerosol generating procedures and the risk of operating room staff’s exposure for aerosol particles.

## INTRODUCTION

Airborne transmission has recently been recognized as an important transmission route of COVID-19 as well as for many other respiratory infections such as tuberculosis and influenza [1-4]. Aerosol particles, which can potentially carry pathogens, are generated for example when breathing, talking, and coughing, but also presumably during certain surgical procedures performed in the respiratory tract area where infective pathogens are known to be found. Fear about the spread of COVID-19 during surgeries has led, for example in the field of otorhinolaryngology, to cancellations of surgeries, minimization of staff in the operating room (OR), and changes in personal protective equipment (PPE) guidelines during the current pandemic [5-7].

Traditionally, aerosols have been defined as particles smaller than 5µm in size that can remain in the air for long periods of time and spread far into space. However, environmental factors have a significant effect on the dispersion of aerosols and there has been a recent debate regarding whether all particles below 100µm should be considered as aerosols [8, 9]. Overall, aerosol generation is a continuum from the smallest aerosols to larger droplet-sized particles, depending on source. Nonetheless, smaller aerosols are present in higher number and tend to spread the farthest and particles smaller than 5µm have been found to carry the majority of pathogens [10-12]. Findings related to COVID-19 support this assumption: SARS-CoV-2 has been detected in particles 0.25– 1.0 μm, 1–4 μm, and > 4 μm. Most results are obtained by PCR, but infectious virus has also been detected in viral culture [13-15]. These facts together make small (<5 µm) particles most challenging for infection control measures[16].

When assessing the risk of infection, the infectious dose associated with the pathogen, the time of exposure and the number of pathogens should be considered. However, both infectious doses of different airborne pathogens and the number of infectious pathogens contained in aerosol particles are yet widely unknown and require further investigation before they can be reliably used as part of a risk assessment for airborne diseases. Therefore, procedures performed in health care, such as surgeries, have been classified as potentially aerosol-generating procedures (AGP) or high-risk AGPs based mainly on the area to be operated and the instruments used, with no degree of quantification [17, 18].

Coughing is known to produce a potentially infectious amount of aerosols and it has recently been used as a quantitative reference for high-risk aerosol generation during surgeries and other clinical procedures [19-21]. It could be said that the current list of aerosol generating procedures is not valid as knowledge of aerosol generation has changed during the pandemic. Many questions persist about quanta and epidemiology in each disease. Procedures in which aerosol generation exceeds the amount of aerosol concentration produced by coughing are those that could be considered t high-risk aerosol generating procedures (AGP) until further knowledge is obtained [22]. This definition does not take a position on the infectious dose, but together with the exposure time provides an estimate of the potential exposure to airborne pathogens

The amount of exposure received by OR staff during the procedure is a key factor in aerosol risk assessment, rather than the total aerosol concentration generated, as highly effective ventilation does lower exposure. Therefore, the purpose of our study was to determine an adequate and quantitative value for high-risk AGPs from the perspective of OR staff by measuring the amount of particle concentration that a person in the OR is exposed to. The results of this study can be applied by comparing the concentration and size distribution of aerosol produced during surgical procedures with the concentration produced by coughing and by estimating the duration of the operation. In the future, as information on infectious doses of various airborne diseases increases, this can be combined with risk assessment.

## METHODS

We recruited 37 healthy volunteers to cough in OR and measured the particle generation during coughing. In addition, 15 patients’ involuntary cough episodes were measured separately when they woke up from general anesthesia or were under local anesthesia. Coughs during extubations were excluded. We compared volitional and involuntary coughing to ensure that there was no significant difference between these allowing a more accurate quantitative assessment of volitional coughs. All measurements were conducted in the Helsinki University Hospital, between December 2020 and February 2021. The ORs had Recair 4C ventilation system with HEPA-14 filtration, and ultra-clean ventilation in the laminar area of 1210–1298 l/s generating 400 - 572,83 air changes per hour. Measurements were performed with the Optical particle sizer (OPS), model TSI model 3330 measuring the size range from 0.3 to 10 µm and flow rate of 1l/min.

OPS was situated 40 cm, 70 cm, and 100cm from the volunteers to reflect the same distances and thus same particle amounts which OR staff, operating physicians, or assisting nurses are exposed to during surgical operations. Involuntary cough measurements were performed an average of 124 cm (range 40–180 cm) from the patient. No additional collection methods, for example funnels, were used to reflect the actual particle exposure in a certain spot (OR personnels’ mouth) in the OR environment. Volunteers were asked to cough as hard as possible for three to five times from each distance. Each cough episode was measured separately ensuring that the particles from previous coughs had time to clear from the OR. Coughs were directed towards the OPS device, and particle concentration was measured with a 5- or 10-second scale interval. For each cough, several measurements were collected for 10 seconds.

The size dependent aerosol concentrations measured with OPS were normalized with respect to the sizing bin widths within 0.3 to 10 µm. The particle number size distributions and total particle concentrations per cubic centimeter were calculated. The particles were categorized based on the diameter as follows: <1 µm, 1–5 µm, and >5 µm. Due to infection risk being related to cumulative aerosol exposure, the mean was calculated for each patient at each coughing distance as a statistical representative. Pairwise comparisons between voluntary and involuntary coughing were calculated using unpaired Student’s t-test with Benjamini-Yekutieli procedure for multiple comparisons with 5% false discovery rate[23]. Prior comparisons the data was log10 normalized. The analyses were performed using Microsoft Excel 2016 (Microsoft Corporation, Redmond, Washington, USA), and GraphPad Prism version 9.0.2 for Mac (GraphPad Software, San Diego, California USA) or RStudio version 1.3.959 (R Foundation for Statistical Computing, Vienna, Austria). Measured minimum concentration in all size classes was 0.000. A p-value <0.05 was considered to be statistically significant. STROBE reporting guidelines were followed when preparing the manuscript.

All procedures that involved human participants were conducted in accordance with the ethical standards of the institutional or national research committee and the 1964 Declaration of Helsinki and its later amendments or comparable ethical standards. The Ethics Committee of Helsinki University Hospital approved the study protocol (HUS/1701/2020). All participants provided written informed consent prior to their participation.

## RESULTS AND ANALYSIS

A total of 306 coughs were measured from 37 healthy volunteers. The detailed information about particle concentrations when coughing from different distances are presented in **Table 1**. All background concentrations were very low (maximum mean total concentration 0.0053 particles/cm³) which enabled the accurate evaluation of particle concentration generated during the procedure. The comparison of involuntary coughs from 15 patients to volitional coughs are presented in **Figure 1**. Mean particle concentration during involuntary coughs was 0.140 p/cm³ ± 0.332 (range 0.006–1.308) for particles <1 µm, 0.025 p/cm³ ± 0.068 (range 0.000–0.270) for particles 1–5 µm and 0.002 p/cm³ ± 0.006 (range 0.000–0.024) for particles >5 µm. These measurements were compared with volitional coughs to determine whether the collected data also describes involuntary coughing. There were no significant differences between volitional and involuntary coughing at any particle size category (p=0.244–0.883).

**Table 1.** Observed particle concentration during volitional coughing from different distances

| Measured cough episodes, n |  | All<br>(n=74) | 40 cm from<br>source<br>(n=37) | 70 cm from<br>source<br>(n=15) | 100 cm from<br>source<br>(n=22) |
| --- | --- | --- | --- | --- | --- |
| Total particle<br>concentration<br>(particles/cm<sup>3</sup>) | Median | 0.055 | 0.055 | 0.026 | 0.070 |
| | Mean ±<br>SD | $1.706 \pm 10.802$ | $0.923 \pm 5.078$ | $0.091 \pm 0.136$ | $4.12 \pm 18.771$ |
|  | Range | 0.000 – 88.157 | 0.013 – 30.973 | 0.000 – 0.466 | 0.016 – 88.157 |
| <1$\mu\text{m}$ particle<br>concentration<br>(particles/cm<sup>3</sup>) | Median | 0.044 | 0.043 | 0.026 | 0.053 |
| | Mean ±<br>SD | $1.692 \pm 10.791$ | $0.906 \pm 5.030$ | $0.087 \pm 0.133$ | $4.111 \pm 18.770$ |
|  | Range | 0.000 – 88.141 | 0.011 – 30.668 | 0.000 – 0.454 | 0.011 – 88.141 |
| 1–5$\mu\text{m}$ particle<br>concentration<br>(particles/cm<sup>3</sup>) | Median | 0.008 | 0.008 | 0.002 | 0.013 |
| | Mean ±<br>SD | $0.013 \pm 0.035$ | $0.017 \pm 0.049$ | $0.004 \pm 0.004$ | $0.013 \pm 0.006$ |
|  | Range | 0.000 – 0.304 | 0.000 – 0.304 | 0.000 – 0.012 | 0.004 – 0.028 |
| >5$\mu\text{m}$ particle<br>concentration<br>(particles/cm<sup>3</sup>) | Median | 0.000 | 0.000 | | 0.001 |
| | Mean ±<br>SD | $0.001 \pm 0.001$ | $0.001 \pm 0.001$ | NA | $0.001 \pm 0.001$ |
|  | Range | 0.000 – 0.005 | 0.000 – 0.005 |  | 0.000 – 0.004 |

SD, standard deviation.

**Figure 1:**
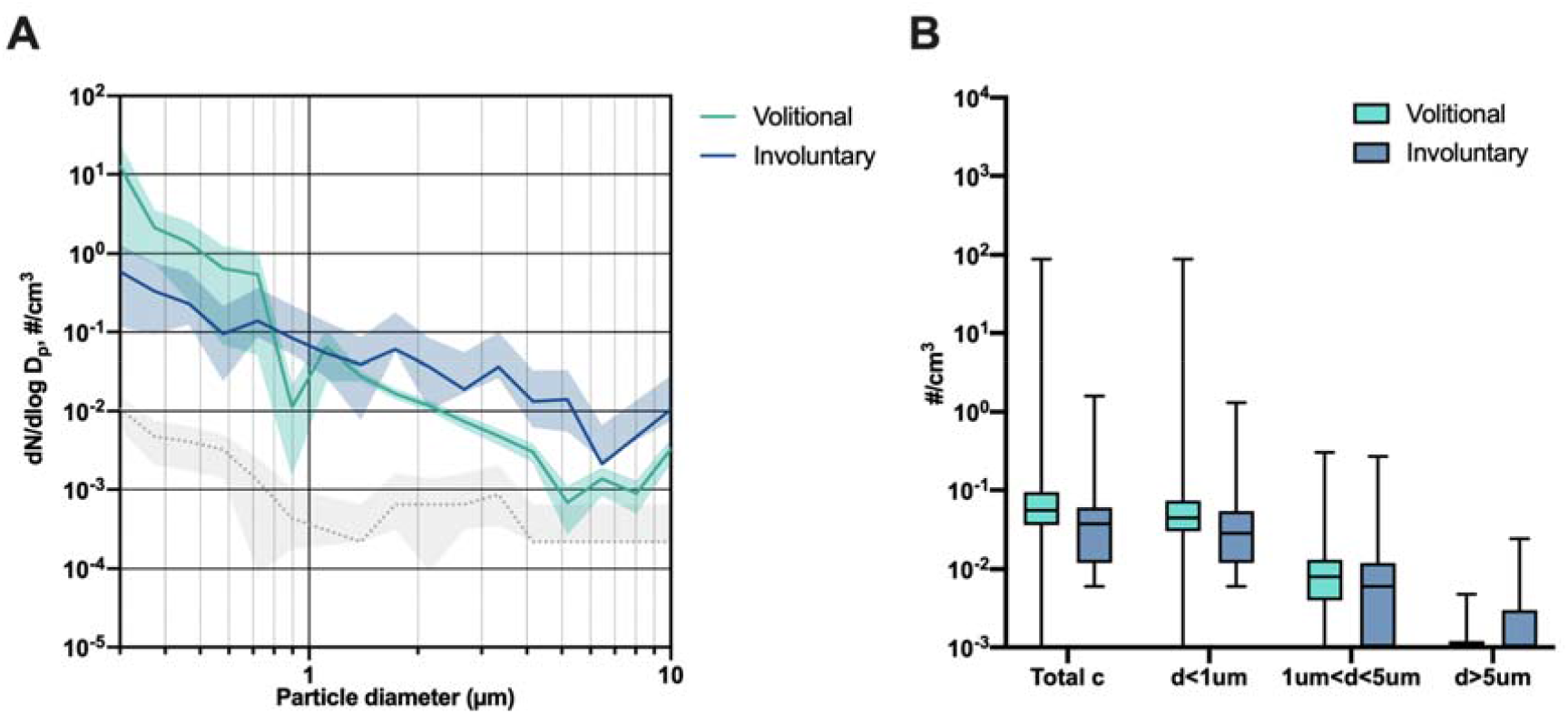
Volitional coughing vs. involuntary coughing. **(A)** Average aerosol size distributions presented with the background (dotted line) during volitional and anesthesia coughs (involuntary) expressed as mean (line) with 95% confidence interval (envelopes). **(B)** Total concentrations and concentrations of <1µm, 1–5µm and >5µm aerosols during volitional and anesthesia coughs (involuntary), presented as median with interquartile range (box) and range (whiskers). Volitionally coughing patients, n=37 (coughs n=306); involuntary coughing patients, n=15 (coughs n=15). c, concentration; Dp, particle diameter; N, particle number

## DISCUSSION

In this study we defined a quantitative estimate for high-risk AGP when assessing the risk of exposure of OR staff. By considering the time spent on the OR, as well as the risk of the patient’s possible airborne disease, a rough risk assessment of the staff’s risk during the procedure was formed. As information on the pathogens contained in aerosol particles as well as infectious doses of airborne diseases increases, these factors can later replace the role of coughing in the risk assessment. Our methodology provides a numeric limit value for the exposure faced by a staff member in an OR environment already considering the environmental factors that affect aerosol dispersion in the space. Thus, the values we measured can be used as a reference when assessing other AGPs that OR staff are exposed to [24].

Our results in relation to the aerosol concentration generated by coughing and the role of coughing as an aerosol generating behaviors (AGBs) are in a line with a recent systematic review. However, we have a higher number of records, we made the systematic distance-dependent evaluation, and we systematically determined aerosol concentrations, which collectively strengthen our study [25]. In our study, the highest aerosol concentrations were observed at 100 cm distance. This is probably due to the methodology: the flow rate of OPS is 1l/min. When measuring particles with high acceleration at close range, some particles bypass the device and are not recorded. When distance increases further, the acceleration of the particles is reduced, and they are observed more accurately. However, OPS is currently the most suitable and used measuring device, especially in OR conditions, so when examining the generation of aerosols during surgical procedures its use is justified despite the limitations.

The large range and personal differences of the particle concentrations observed in our study is naturally seen in different respiratory activities and related to heterogeneity of the cough[26]. However, taking into account similar concentrations of volitional and involuntary coughs, we can state that the presented data is well representative of the average aerosol concentrations generated during coughing. A previous study of Lee et al. showed that infected patients generated a greater number of particles when coughing compared to the healthy ones[27]. Thus, it could be stated that the particle concentration seen in our results is the minimum value to determine the limit of AGP.

Whether the definition for AGP is useful at all can be discussed. Understanding humans as aerosol generators during normal respiratory activities has grown and the term aerosol generating behaviors (AGBs) are proposed alongside the AGP [25] However, statistics from around the world show that surgeries involving mucus membranes and respiratory track area were significantly reversed during the COVID-19 pandemic due to the fear of infection. Thus, quantitative variables used for risk assessment are a step forward from previous intuitive measures toward more comprehensive risk assessment.

## CONCLUSION

This study provides a standard for aerosol concentration and size distribution for aerosols generated when coughing to act as reference for high-risk aerosol generation during surgical procedures performed in the OR.

## Data Availability

The authors confirm that the data supporting the findings of this study are available within the article [and/or] its supplementary materials.

## ABBREVIATIONS

(AGP): Aerosol generating procedure
(OR): operating room

## ACKNOWLEDGMENTS

We thank Catharina Pomoell for her great work as a research nurse

## INFORMED CONSENT

All patients provided written informed consent prior to participating in the study.

## ROLES OF AUTHORS BASED ON ICJME STANDARDS

All authors have approved the manuscript and have made significant contributions:

Conceptualization (ES, LMO, VJA, LL); methodology (ES, LMO, APH, VJA, AG); validation (ES, LMO, APH); formal analysis (ES, LOM, NR, ML, APH); investigation (ES, LMO, NR); resources (ES, AG); writing original draft (ES); writing – review & editing (LMO, NR, ML, APH, LL, VJA, AG); visualization (NR, ML); editing (ES); project administration (ES, LMO, AG; VJA); funding (ES; APH; AG)

